# Association Analysis of *CYP2A6* Gene Variant (rs1801272A>T) with Nicotine Metabolism and Smoking Tendency Among Pakistani Youth

**DOI:** 10.1101/2025.04.16.25325925

**Authors:** Iqra Yasmin, Haider Ali, Muhammad Rafeh, Muhammad Sikandar, Abdul Kashif, Muhammad Salahuldin, Ammad Shafeeq, Rashid Saif

## Abstract

**Background:** Cytochrome P450 2A6 (CYP2A6) is a key enzyme in nicotine metabolism, with its genetic variants playing a role in smoking behavior. Particularly, g.40848628A>T is significantly associated with nicotine metabolism and smoking tendency in different populations. The purpose of the current study is to examine the genetic diversity of this locus and association analysis within smokers and non-smokers cohorts among Pakistani youth.

**Methods and Results:** The allele-specific ARMS PCR genotyping technique was applied to examine a total of 100 samples as a case-control study of n=50 from each cohort. From the sampled individuals, 92% were found to be homozygous wild-type (AA), 7% were heterozygous (AT), and 1% were homozygous mutant (TT). PLINK software was used for the Chi-square test yielded, χ^2^ (1, N = 100) = 2.91, p = .088 suggesting a non-significant trend towards association, where alternative allele frequencies were calculated as 0.07 and 0.02 in cases & control cohorts respectively. Similarly, Hardy-Weinberg Equilibrium (HWE) p = 0.1714 indicates genotype frequencies did not significantly deviate from HW expectations and no error or selection in the overall samples. The carriers of the alternative allele have 3.688 times higher odds of being affected by the condition compared to non-carriers with the reference allele.

**Conclusions:** Future studies with larger sample size may help to clarify the population structure of the subject locus. Genome-wide association studies using next-generation sequencing may also aid in predicting nicotine metabolism and resistance to smoking cessation in the Pakistani population.

## Introduction

Cigarette smoking, a prevalent source of nicotine, poses significant health risks across all age groups, particularly with a concerning surge in popularity among Pakistani adolescents. Nicotine, a proven addictive substance, exhibits varying degrees of dependence in individuals due to the metabolism by the Cytochrome P450 family 2 subfamily A member 6 (CYP2A6) gene product [1, 2]. This primary protein transforms nicotine into its byproduct cotinine and other metabolites. Divergent levels of CYP2A6 protein influence nicotine metabolism, impacting an individual’s tendency towards smoking. Wild-type CYP2A6 proteins with normal activity facilitate the standard breakdown of nicotine, mitigating its effects on the brain and discouraging the development of smoking dependence. Conversely, mutated CYP2A6 proteins with impaired activity hinder nicotine metabolism, allowing its retention in the body, leading to prolonged brain effects with relatively lower cigarette consumption [3, 4].

Despite the well-known health risks associated with smoking, it remains a substantial global health burden. Adult smoking prevalence is 32.6% and 6.5% in men and women respectively, contributing to ∼7.7 million annual deaths worldwide among its total smokers of 1.14 billion [5]. Alarmingly, low and middle-income countries, constituting 80% of total smokers globally, face an increased prevalence of tobacco use [6]. In Pakistan, 13.4% age-standardized prevalence of tobacco use is reported in urban and rural areas with its alarming rates of 16.3% & 11.7% respectively [7]. This escalating trend of tobacco use in Pakistan has prompted the scientific community to delve into its genetic aspects involved in smoking tendency, levels of cigarette consumption, depth of inhalation and smoking cessation ability of the individuals [7-9].

The *CYP2A6* gene exhibits numerous variants associated with nicotine metabolism across diverse populations [10]. The subject variant located on Chr.19 NC_000019.10 at 19q13.2 locus (NG_008377.1: g.6820T>A, & (ATG start) position 1799T>A). Chromosomal positioning is g.40848628A>T in the (GCF_000001405.40) genome assembly, current variant corresponds to c.479T>A (r.500) in the NM_000762.6 transcript, affecting the protein’s p.L160H position, situated on exon 3 of total 6907 nucleotide position of *CYP2A6* gene. This variant impacts the protein’s function, reflected in its encoded protein ID NP_000753.3 of 494 amino acids. Notably, rs1801272A>T, associated with poor nicotine metabolism, has a global AAF (T=0.0092) as per the 1000Genome database [11]. The (A) allele signifies normal CYP2A6 protein activity, while the (T) allele may alter activity, influencing nicotine metabolism [12]. The subject variant was genotyped through allele-specific ARMS-PCR, followed by assessments for (HWE), (χ2), and (OR) statistics. Subsequently, the derived (AAF) will be determined in both cases and controls. These quantitative analyses aim to calculate association and correlation coefficients, providing valuable insights into the relationship between the variant, nicotine metabolism and smoking tendency among Pakistani youth.

## Materials and Methods

### Sample collection and DNA extraction

A comprehensive genotyping study of 100 blood samples, encompassing specimens from both adolescent/youth smokers (n=50) and non-smokers controls (n=50). The primary objective is to investigate the potential association and correlation between the *CYP2A6* locus with nicotine metabolism/dependency among Pakistani youth. Blood samples from male adolescents ranging 18-30 years were collected using K3-EDTA vacutainers and stored at 4°C until subsequent analysis. Genomic DNA extraction was performed using column-based kit method (<u>www.favorgen.com</u>), adhering to the manufacturer’s instructions for precise and consistent DNA extraction.

### Primer designing

The allele-specific ARMS primers were meticulously designed through specialized software such as OligoCalc and NetPrimer. The ARMS-PCR protocol was employed to amplify both wild and mutant-type variants from each sample. Specifically, one ARMS primer was designed for each variant, strategically incorporating a mismatch at the 4nt position from the 3’ end of the sequence. Additionally, to uphold PCR accuracy, two internal control (IC) primers were also employed in the experimental setup during optimization. This comprehensive primer design (Table 1) and PCR strategy aim to enhance the precision and reliability of the genetic variant analysis [13].

**Table 1.**
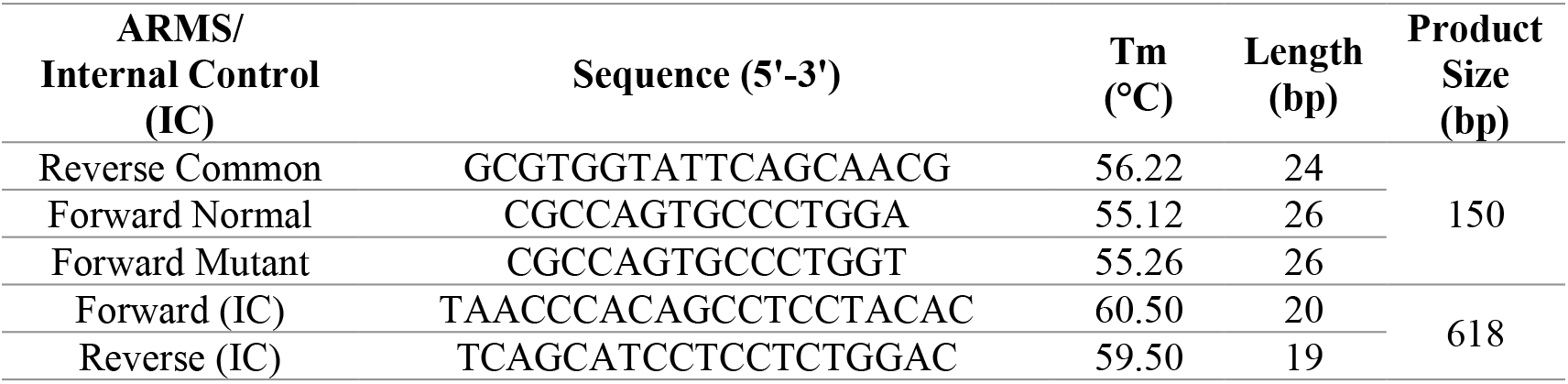
Primers sequence attributes.

### PCR amplification

Each sample, encompassing both wild-type and mutant-type variants were undergoing amplification using a thermocycler. Two distinct PCR reactions were executed for each sample, with each reaction involving the reverse common primer paired separately with forward ARMS primers designed for the wild and mutant-type allele. Concurrently, the forward and reverse internal control (IC) primers were also utilized to amplify internal control regions during optimization process. A reaction mixture of 14μL was prepared, comprising 1μL of 50ng/μL genomic DNA, 10mM of each primer, 0.015 IU/μL of *Taq* polymerase, 2.5mM of each MgCl2 & dNTPs, 1x Taq buffer along with PCR-grade water DEPC treated water. The touch-down PCR protocol was initiated with an initial denaturation at 94°C for 3 minutes, followed by 35 cycles of denaturation at 94°C for 30 seconds, annealing started from 58.4°C with decrease of 0.5°C/cycle for 10 cycles and remaining 25 cycles were executed at 54°C for 30 seconds, extension at 72°C for 30 seconds, concluding with a final extension at 72°C for 07 minutes [14]. Subsequently, the reaction mixture was stored at 4°C. This detailed PCR procedure aims to ensure precise and robust amplification of target regions in the genomic DNA (Figure 1).

**Figure 1:**
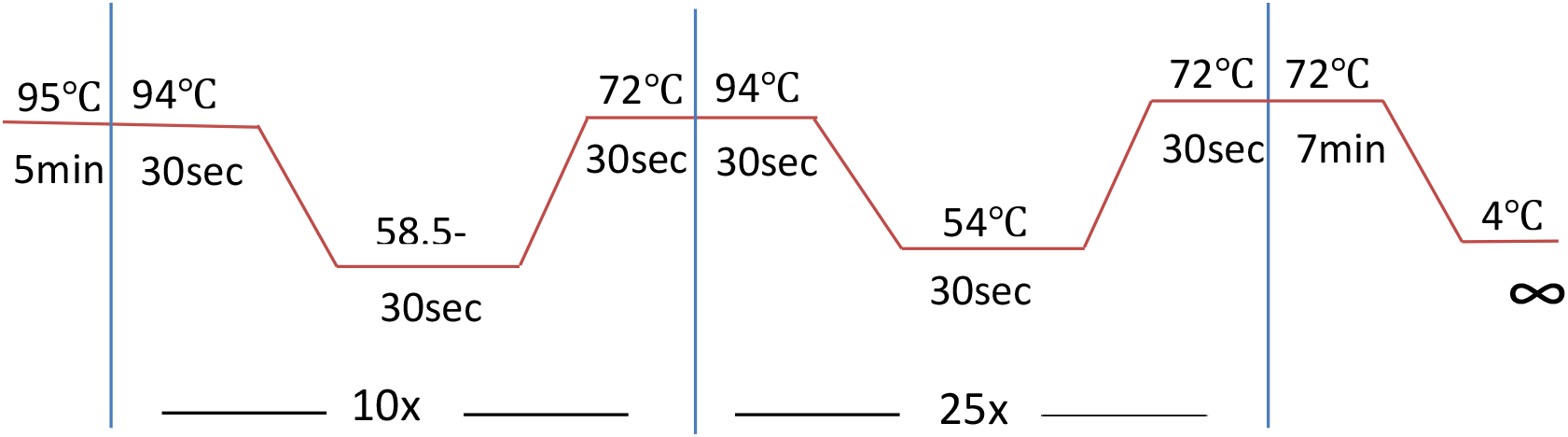
Thermal cyclic conditions of ARMS-PCR

### Statistical analysis

The PLINK data analysis toolset was used in computing both observed and expected genotyping frequencies, incorporating considerations for HWE through the application of the equation *p*2 + 2*pq* + *q*2 = 1. The analysis was extended to Chi-square testing, employing the formula 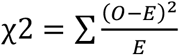 to ascertain the association between the subject variant rs1801272 with nicotine metabolism/dependency and smoking behavior within the sample set. Additionally, *p-values* and odds-ratios (OR) were undertaken followed by alternative allele frequencies for further enriching the statistical insights.

## Results

In the current study, *CYP2A6* gene variant (rs1801272A>T) showed the variability in Pakistani sampled population, which is in accordance to the previous studies and other world populations, and depicts Chr.19 locus g.40848628 is variable and under-selection as showed in (Figure 2).

**Figure 2:**
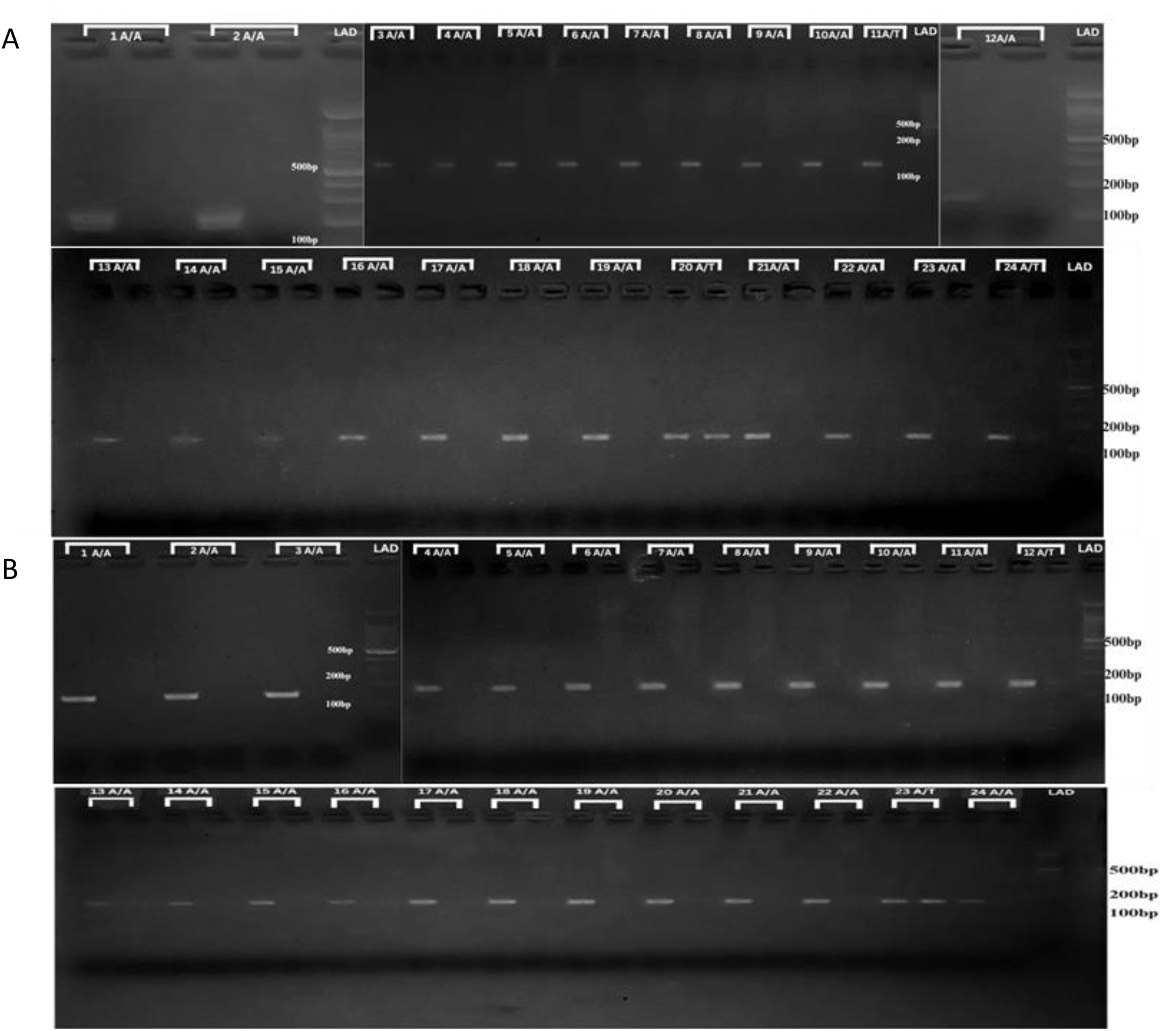
Gel picture of ARMS-PCR amplification of targeted variant within cases A) and controls B)

A total of 100 samples were genotyped, (smokers=50) and (non-controls=50). After experimental and statistical analyses, it was concluded that within smoker’s cohort, there are 05 heterozygous (AT), 44 homozygous-wild (AA) and 01 sample was observed as homozygous-mutant. Similarly, within controls 02, 48 individuals are heterozygous, homozygous-wild and none of the sample was observed as homozygous-mutant respectively. Remaining samples gel pictures are provided in *Supplementary Fig. 1*. Overall genotypic frequency of homozygous-wild in our sampled population is 0.92, heterozygous are 0.07 and homozygous-mutant is 0.01. Subsequently, (*HWE*) Chi-square analysis was also conducted to verify, whether our sampled population is obeying this principle or not with the following outcomes of *χ*2(1, *N* = 100), *p* = .1714 which manifests that our sampled population is in accordance with the *HWE* equilibrium as the *p*-value is above the set threshold confidence interval of 0.05 so accepting our null-hypothesis of observing *HWE*, means no selection and population is randomly bred.

Moreover, alternative allele frequencies were also observed as 0.07 & 0.02 within our cases & control cohorts having *χ*2 statistics value of 2.90 and *p* = 0.088 showing no significant association of the screened variant with nicotine metabolism and smoking tendency in Pakistani youth. Likewise, Odds-ratio (OR) of 3.688 was also calculated showing the prevalence of odds/mutant variant is ∼4 times higher in cases vs controls, depicts the higher relative risk of subject phenotype. (Table 2).

**Table 2.** Plink association of *CYP2A6* gene variant (rs1801272A>T) with nicotine metabolism and smoking tendency among Pakistani youth.

| Sam<br>ple # | Chr.<br>Pos. | cDNA variant<br>NM_000762.6 | Protein<br>variant<br>NP_00075<br>3.3 | Genotypic<br>Frequencies |  |  | Alternative<br>Allele<br>Frequencies |  | *p*-value<br>(OR) |
| --- | --- | --- | --- | --- | --- | --- | --- | --- | --- |
|  |  |  |  | Homo-<br>wild<br>(AA)% | Hetero-<br>(AT)% | Homo-<br>mutant<br>(TT)% | Case | Contr. |  |
| 100 | 19:4084862<br>8 | c.479A>T<br>r.500 | p. L160H | 92 | 7 | 1 | 0.07 | 0.02 | 0.088<br>(3.68) |

As the individuals with “ T” allele tend to metabolize nicotine more slowly, leading to lower cigarette consumption and potentially reduced risk of nicotine addiction [15, 16]. Subject cDNA variant c.479A>T alters lucine amino acid (aa) to histidine at 160^th^ position, and this (aa) is responsible for slower nicotine metabolism that may find it easier to quit smoking compared to those with normal *CYP2A6* activity. Further, functional genetics studies are still needed to confirm and validate this postulated hypothesis.

Another statistical test, Cochran-Armitage Trend test was also conducted to evaluate the association between subject variant and our trait, assessing whether the frequency of minor allele differs between affected and unaffected individuals. The test yielded a chi-square value of 2.453 (df = 1) and a *p* − *value* = 0.1173, indicating no significant association, which suggests that the genotype distribution at this variant does not show a meaningful trend between allele dosage and disease status in our sampled population.

Similarly, dominant (DOM) and recessive (REC) models testing were also tested to explore the potential genetic association of our subject variant with the nicotine metabolism. However, both models yielded NA values for chi-square and *p* − *values*, indicating lack of variation in genotype distribution to perform statistical testing. This suggests that neither a dominant nor a recessive model pattern could be established for this variant in the given dataset, because only (1+5) = 6 vs. 44 genotypes were observed in affected and (0+2) = 2 vs. 48 in unaffected population respectively.

The logistic regression analysis was also applied under the additive genetic model which effectively models binary traits while adjusting covariates suggesting that for each additional copy of the affected allele, the odds of being affected increase by a factor of 3.16. The additive model assumes a linear effect of each additional risk allele on subject phenotype susceptibility. However, the *p* − *value* = 0.1455 is not significant, indicating no strong association due to limited sample size, low allele frequency or random variation.

## Discussion

The current study examined the association between the *CYP2A6* gene variant rs1801272A>T with nicotine metabolism and smoking behavior among Pakistani youth. The statistics revealed no significant association with nicotine metabolism and smoking tendency among our sampled population. Current results showed overall 92% of the sampled population was homozygous-wild (AA), 7% was heterozygous (AT), 1% was homozygous-mutant (TT) with *p* = 0.088, and *OR* = 3.68.

Prior research around the globe has reported mixed findings on *CYP2A6* polymorphisms in nicotine metabolism. Studies on European populations indicate that the rs1801272(A) allele frequency is around 5% in CEU and IBS populations, with a reported *p* = 0.81 [17]. In the Mexican population, the allele frequency was found to be <1%, leading to its exclusion from statistical analysis due to low prevalence [17]. An Egyptian population study reported an association between this variant and nicotine metabolism variation, with statistical significance varying across studies *p* = 0.27 [18]. In contrast, Japanese populations exhibit a higher frequency with reduced function *CYP2A6* protein, correlating with slower nicotine metabolism and altered smoking tendencies, with a reported *p* = *value* 0.034 [19]. Our genotypic distribution showed that among cases, only five individuals were heterozygous (AT), while the majority (44/50) were homozygous wild-type (AA). In controls, two individuals carried the heterozygous variant, with the rest (48/50) being homozygous-wild. This suggests that rs1801272T is rare in the Pakistani population, limiting its influence on smoking behavior [1].

Nicotine metabolism is primarily mediated by *CYP2A6*, which converts nicotine into cotinine. Genetic variations in *CYP2A6* may influence smoking initiation, dependence and cessation activities. Studies indicate that slow metabolizers, due to reduced function of CYP2A6 protein, tend to smoke less and experience lower nicotine dependence [20]. As for as the clinical relevance and pharmacogenomics is concerned this variant may influence the effectiveness of the nicotine replacement therapy (NRT) for smoking cessation [18, 19, 21]. However, our findings suggest that rs1801272A>T alone is not a major determinant of smoking behavior in Pakistani youth.

## Conclusion

This pilot study suggested the variability of the subject CYP2A6 gene locus rs1801272A>T and seems not significantly associated with nicotine metabolism and smoking tendency among Pakistani youth with *p* = 0.088 and *OR* = 3.68. Further, large sample size single locus genotyping or GWA studies may be conducted to evaluate the smoking genetics in Pakistani population.

## Supporting information

Sample collection request & Ethical IRB approvals

## Acknowledgement

Authors are obliged to the Diagnostic Zone Lab-Lahore and Shah Medical Center-Swat for providing blood samples.

## Authors Contribution

Iqra Yasmin (IY), Haider Ali (HA), Muhammad Rafeh (MR), Muhammad Sikander (MS), Abdul Kashif (AK) performed wet-lab (blood collection, DNA extraction, PCR and gel electrophoresis) and wrote the initial draft, Muhammad Salahuldin (MS), Ammad Shafeeq (AS) helped in blood samples collection. Rashid Saif (RS) designed and envisaged the research project, trouble shooting in wet & dry-labs, applied statistical tests, editing, proof reading and correspondence of the manuscript.

## Statement of Ethics

Through proper channel ethical guidelines were followed for sample collection

## AI Tool Declaration

No AI tool was used to prepare this manuscript.

## Data Availability

Complete data of the article is available within the manuscript and supplementary file(s).

