## Supplementary figures and images for "Association Analysis of *CYP2A6* Gene Variant (rs1801272A>T) with Nicotine Metabolism and Smoking Tendency Among Pakistani Youth"

### Supplementary Fig.1

Supplementary Figure 1. Showing cases and controls genotyping results

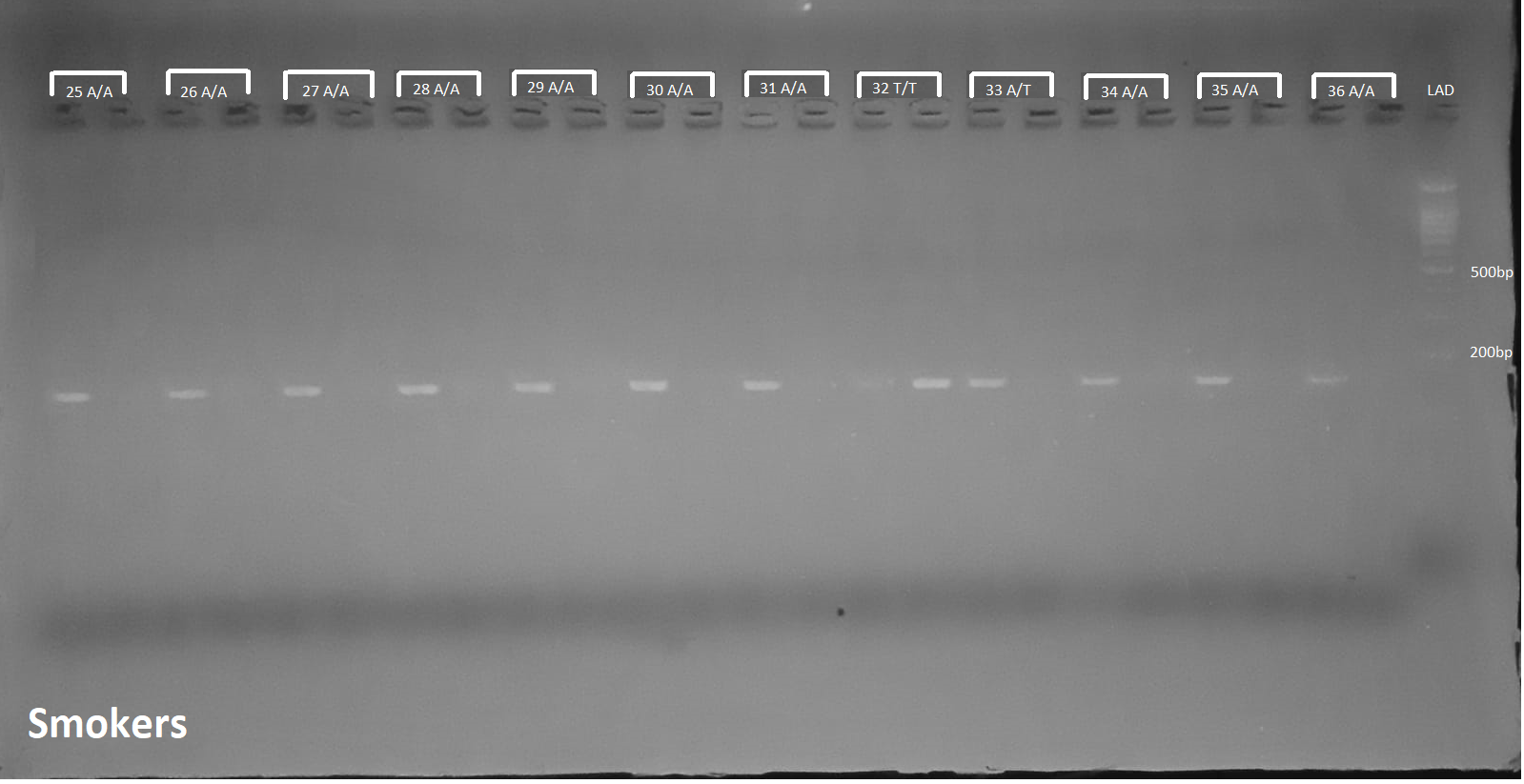

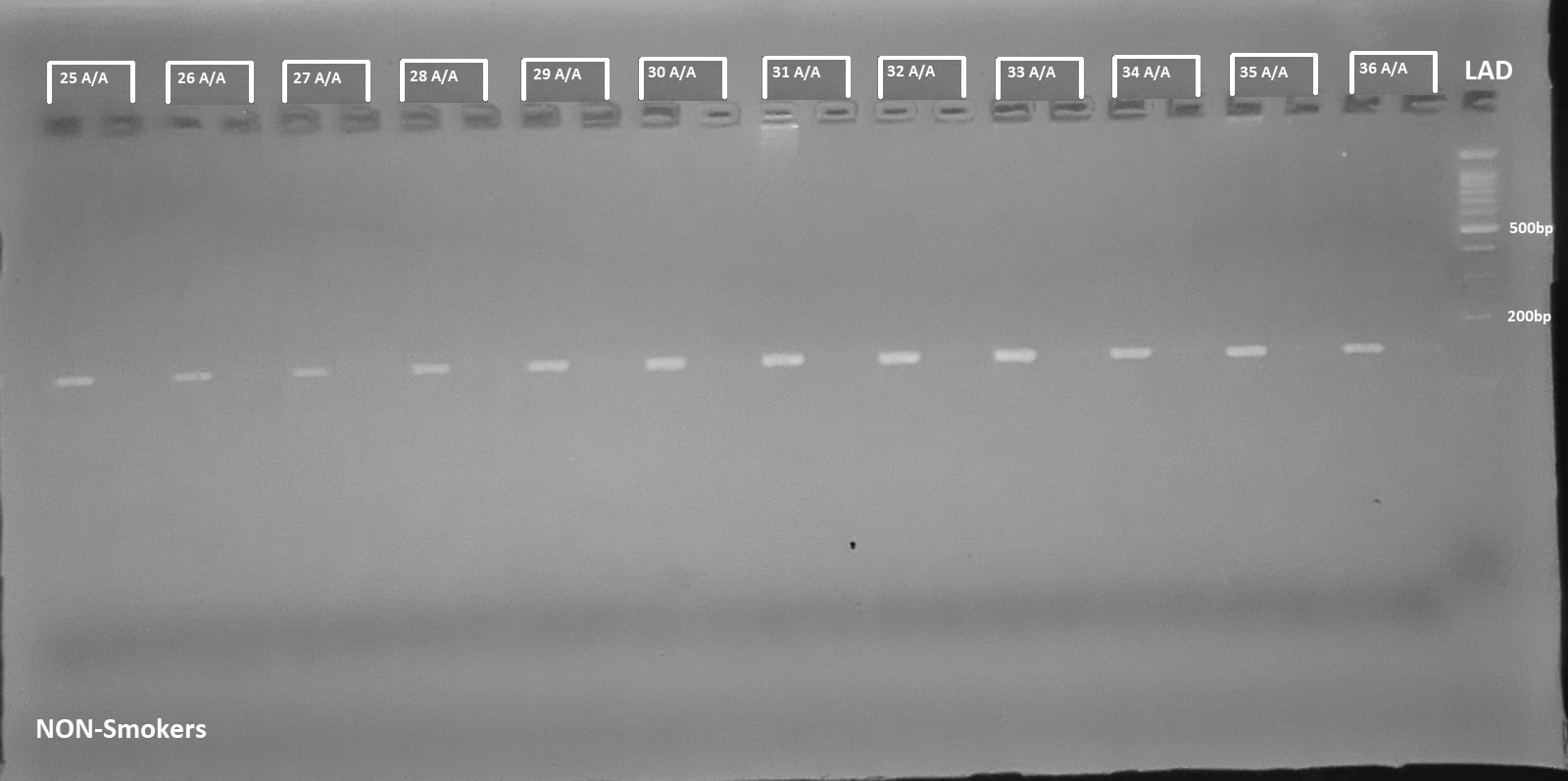

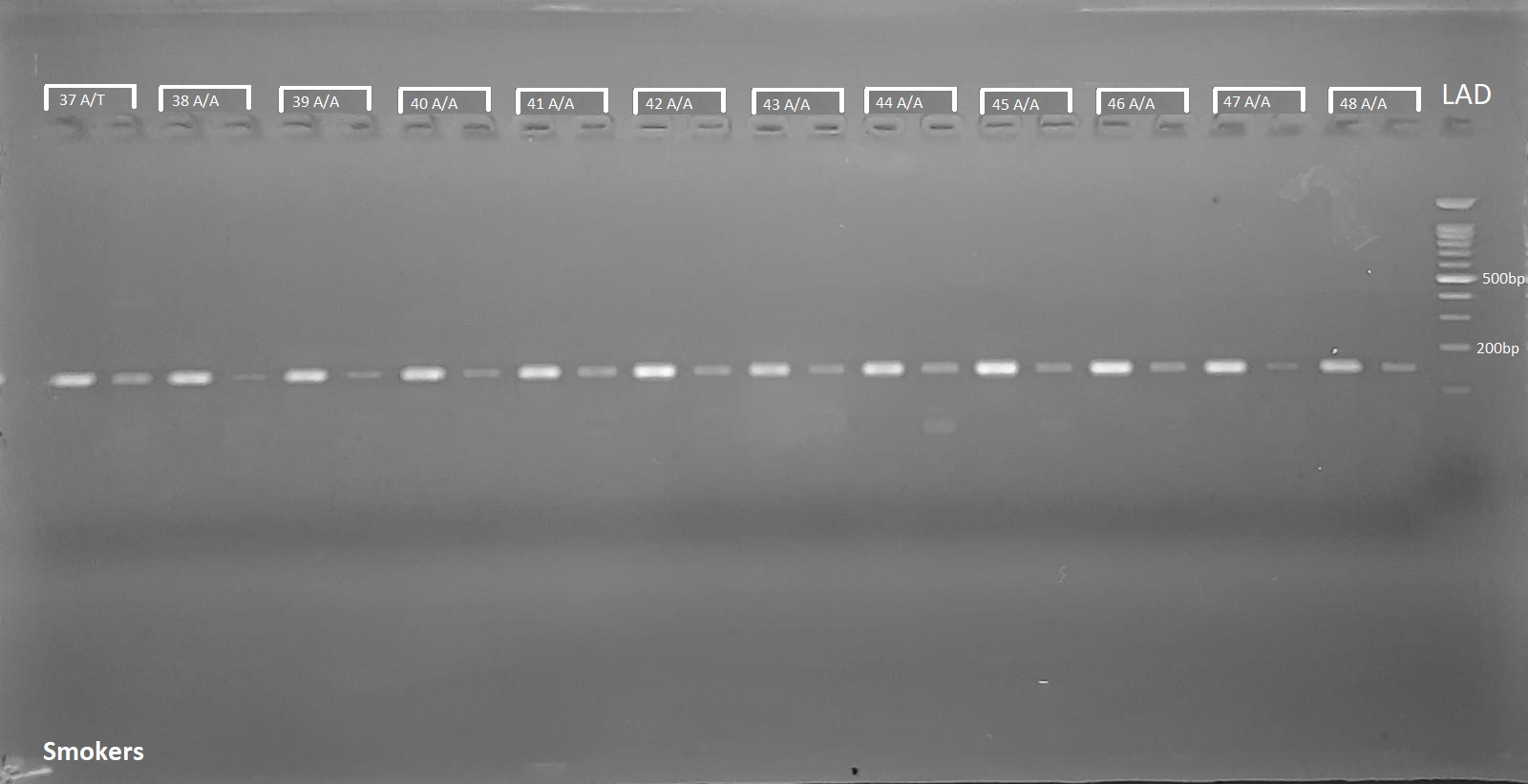

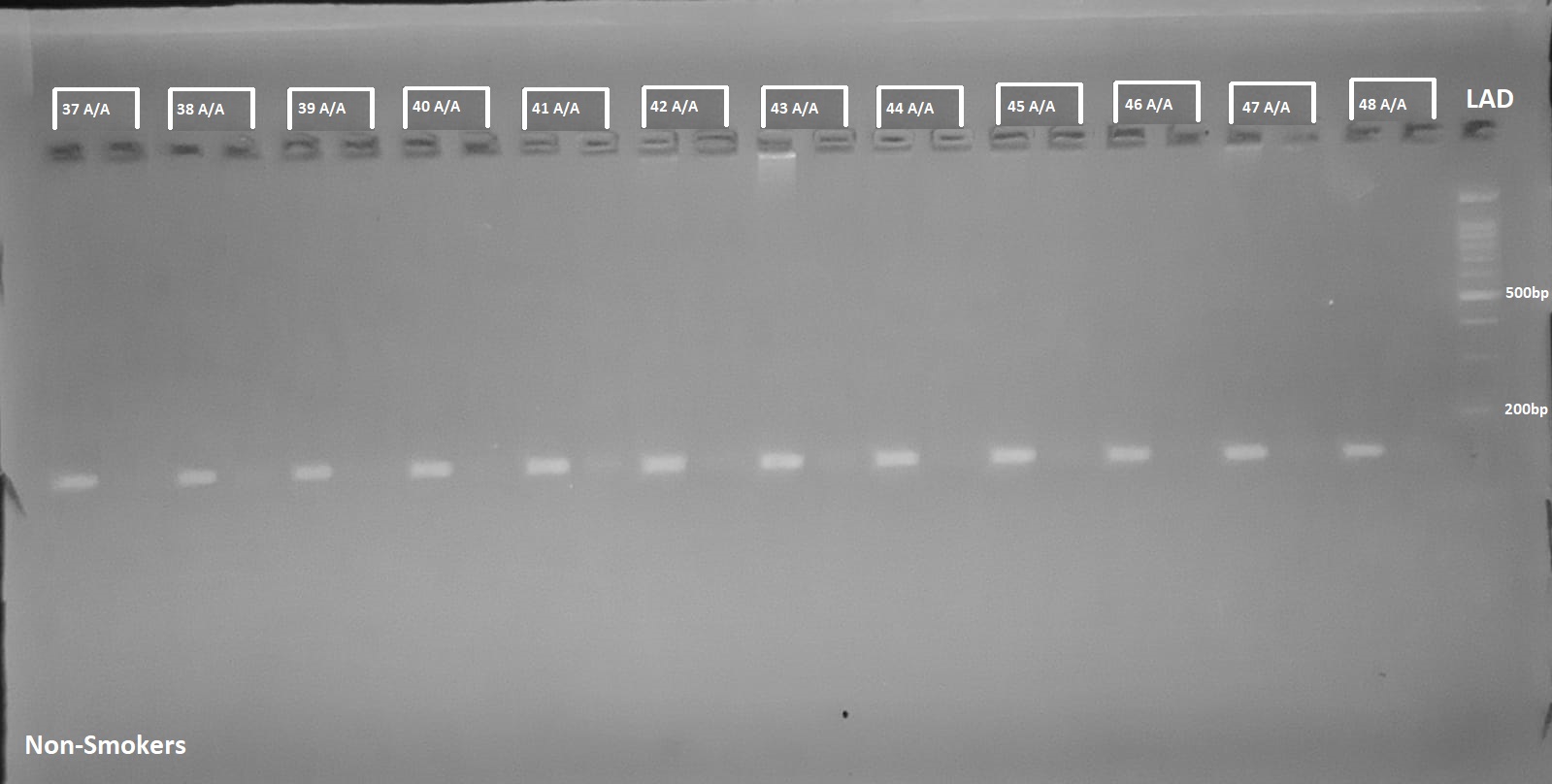
