## Supplementary material for "Association Analysis of *CYP2A6* Gene Variant (rs1801272A>T) with Nicotine Metabolism and Smoking Tendency Among Pakistani Youth": Sample collection request & Ethical IRB approvals

To
The Lab Manager

**Subject: Request for Provision of Blood Samples**

Dear Sir,

I hope this message finds you well. I am reaching out on behalf of a research initiative focused on understanding the human genetics of nicotine dependency in Pakistani youth. This team is composed of five BS-Biotechnology students at Qarshi University mentioned below.

We have learned about the valuable contribution of your esteemed Lab in the health sector, and we believe that collaborating with your institution would significantly contribute to the success of our research. With this in mind, I kindly request your support in providing blood samples from individuals within your lab for our study, and subsequently, your institution's name will be acknowledged in the publication.

The data collected will be treated with the utmost confidentiality and used exclusively to advance our understanding of nicotine dependency. Our goal is to contribute to the development of more effective molecular diagnostics and genetic association studies for this global health challenge.

If you have any questions or require additional information, please feel free to contact me at the below signatures. I appreciate your consideration of this request and look forward to the possibility of working together to make meaningful strides in the field of smoking behavior and nicotine intake research.

Thank you for your consideration.

Sincerely,

Dr. Rashid Saif, *PhD, Postdoc (Switzerland)*
Associate Professor | HoD-Biotechnology | Head ORIC
Qarshi University, Lahore

Students names:
Haider Ali
Muhammad Sikanadar
Muhammad Rafeh
Iqra Yasmeen
Abdul Kashif

Reference No.: DZL#004/25

**To Whom It May Concern**

This is to confirm that Diagnostic Zone Lab has actively collaborated in the research project entitled: **"Association Analysis of CYP2A6 Gene Variant (rs1801272A>T) with Nicotine Metabolism and Smoking Tendency Among Pakistani Youth."**

As part of this collaboration, Diagnostic Zone Lab facilitated the collection of blood samples in full compliance with the approved Standard Operating Procedures (SOPs). All samples were collected by trained phlebotomists while strictly adhering to national and international ethical, biosafety, and bioethical standards. The lab ensured that the privacy, confidentiality, and rights of all participants were fully respected and protected throughout the process.

The collaboration also involved maintaining the integrity of the sample collection, handling, and storage procedures in alignment with the project's scientific and ethical requirements.

Should you require any further information or clarification, please feel free to contact our office.

Sincerely,

DIAGNOSTIC ZONE LAB DZL

Diagnostic Zone Lab
Awasia Society, College Rd.,
Lahore, Pakistan

Date: Aug 09, 2024




CS Scanned with CamScanner

SHAH
MEDICAL CENTER

**Reg# 04024-R**

**To whom It May Concern**

This is to certify that SHAH medical center has reviewed and granted ethical clearance for the researc study titled, **"Association Analysis of *CYP2A6* Gene Variant (rs1801272A>T) With Nicotin Metabolism and Smoking Tendency Among Pakistani Youth."**

The investigators submitted all essential documents, including the research methodology and protocols fo sample and data management. These were assessed in line with both national and international ethica standards for studies involving human participants. In addition, blood samples were collected at SHA medical center by trained phlebotomists, in strict compliance with bioethical and biosafety procedures Prior to the collection, written informed consent was obtained from all participants. Participan confidentiality, privacy, and rights were fully protected throughout the process.

For any further inquiries or clarification, please feel free to reach out to our office.

Shah Medical Cente
Opp: Civil hospital Bariko
Ph: 0946-751515

For verification: 0946-751515

Opposite THQ Hospital Barikot Swat Pakista
